# Depression and anxiety among the University community during the Covid-19 pandemic: a study in Southern Brazil

**DOI:** 10.1101/2021.04.12.21255340

**Authors:** Helena S. Schuch, Mariana G. Cademartori, Valesca Doro Dias, Mateus L. Levandowski, Tiago N Munhoz, Pedro R. C. Hallal, Flávio F. Demarco

**Affiliations:** Graduate Program in Dentistry, Federal University of Pelotas; Undergraduate Student, Federal University of Pelotas; School of Psychology, Federal University of Pelotas; Postgraduate Program in Psychology, Federal University of Rio Grande; Postgraduate Program in Epidemiology, Federal University of Pelotas

**Keywords:** COVID-19, Mental Health, Mental Disorders, Depressive Symptoms, Universities, Student Health Services

## Abstract

**Background:** The COVID-19 pandemic has disrupted people’s routine in several ways, including the temporary cessation of face-to-face teaching activities, which may affect the mental health of the population. This study aimed to assess the mental health of the academic community of a University in South Brazil during the Covid-19 pandemic.

**Methods:** Cross-sectional web-based survey conducted between July-August 2020 through a self-administered online questionnaire. All University staff and students were eligible. Depression was assessed using the Patient Health Questionnaire-9 and anxiety by the Generalized Anxiety Disorder-7. All analyses were stratified by academic or administrative staff, undergraduate and graduate students.

**Results:** 2,785 individuals participated in the study. Prevalence of depression and anxiety were 39.2% (95% Confidence Interval (CI) 37.3-41.1) and 52.5% (95% CI 50.6-54.4), respectively. On stratified analyses, undergraduate students showed a higher prevalence of the outcomes compared to other groups. In relation to social distancing, higher prevalence of mental illness was associated to strictly following the authority’s guidelines and with not leaving the house routinely, but these associations were restricted to some subgroups. Mental health care and previous diagnosis of mental illness were associated with higher rates of anxiety and depression.

**Limitations:** The main concerns were the representativeness of the sample and the response rate.

**Conclusions:** An alarming prevalence of mental illness was observed in this academic community. Despite the well-known benefits of social distancing and quarantine to public health, it requires a special surveillance on the mental health of the population, especially students and those with previous mental illness diagnosis.

## Introduction

In March 2020, the World Health Organization declared the COVID-19 outbreak as a pandemic after 118,000 cases and 4,291 deaths reported in 114 countries (World Health Organization, 2020). One year later, in the end of March 2021, more than 125,000,000 cases and almost 2,750,000 deaths have been reported. Brazil is the third leading country in relation to number of cases and the second in relation to deaths, with cases and deaths in the country representing around 12% of the global figures, despite having only 2.7% of the world population (Dataset, 2020).

During an extreme situation such as a pandemic, the focus and efforts of health professionals, scientists and government naturally turn to the biological risks of the disease, seeking to understand the pathophysiological mechanisms and proposing measures to prevent, contain and treat the disease, such as potential medications and vaccines. In this context, the secondary effect, although also highly relevant, on individual and societal mental health, tends to be underestimated and neglected. (Ornell et al., 2020).

The SARS-CoV-2 virus, responsible for COVID-19, is highly infectious in humans and has a worrisome mortality rate. The virus is relatively new, and there is still a lot to be studied about its evolutionary ancestry, the diagnosis of COVID-19 and its treatment. The SARS-CoV-2 virus and the related disease have induced widespread panic and anxiety, due to its still unknown characteristics (Banerjee, 2020), in addition to the well-known high transmissibility of the virus and the recommendations for social distancing.

The COVID-19 pandemic has been associated with anxiety, depression, stress, sleep disorders and suicide (Sher, 2020). In a pandemic, fear increases anxiety and stress levels in healthy individuals and intensifies the symptoms of those with pre-existing psychiatric disorders (Shigemura et al., 2020). In addition to the fear of the disease, the COVID-19 pandemic affects several aspects of individuals’ lives, such as family organization, changes in routine, with the closure of schools, universities and commerce, and the recommendation of social distancing, with possible feelings of abandonment and loneliness. There is also insecurity and fear regarding the socioeconomic implications of the pandemic (Ornell et al., 2020). The United Nations Secretary-General, António Guterres, draws attention to the impacts of the pandemic on the individuals’ mental health, not only during the problem, but also when it is already under control. As highlighted by the Secretary-General, even after the pandemic, mourning, anxiety, and depression related to COVID-19 will continue to affect people and communities (United Nations, 2020).

Since the beginning of the pandemic, University routines were significantly changed. Most of the Universities around the world were closed and teaching and learning process needed to be modified for the remote digital environment (UNESCO, 2020). In Brazil, most Universities ceased in person activities during the entire year of 2020, especially due to the lack of pandemic control, with the country remaining a COVID19 hot spot. University students are characterized as especially vulnerable to the effects of the pandemic on mental health. They constitute a population in transition, entering adult life and experiencing economic and social changes. In addition, with the implementation of a single national examination as the main mean of entry into Brazilian federal institutions (Exame Nacional do Ensino Médio – ENEM), a large proportion of university students move intercity or even interstate to attend a Federal University, which is public and free of charge. Adapting in a new city, often with limited social support, can increase the emotional vulnerability of these students. In fact, the UN identifies adolescents and young people as especially at-risk populations, and recognizes concerns about family health, closure of schools and universities, loss of routine and loss of social connection as the main sources of distress (United Nations, 2020). A recent study with first year students from the Federal University of Pelotas (UFPel) showed that 32% of them had at least one major depressive episode, with it being more frequent among women, who had a family history of depression, who belonged to sexual minorities or who lived with friends and colleagues. Differences on the occurrence of depressive disorders were also observed according to the undergraduate course the students were enrolled at, while a poor academic performance, the abuse of alcohol and illicit drugs were associated with a higher prevalence of mental disorders (Flesch et al., 2020).

Public University Staff have been the subject of studies regarding their mental health. In Brazil, these professionals have job stability and in general have an average salary higher than those who perform the same function in the private sector. They usually perform in person activities. However, due to the COVID-19 pandemic, almost all staff have been carrying out activities remotely (home-office), which can lead to stress and mental disorders.

Considering the potential impacts of COVID-19 on the mental health of the population in general and the increased vulnerability of the University population, it is important to monitor the impact of the pandemic on this group. Therefore, the aim of this study is to assess the mental health of the academic community of a University in South Brazil during the Covid-19 pandemic.

## Materials and methods

### Study site

This study was carried out with the community of the Federal University of Pelotas (UFPel), a public university located in the city of Pelotas, Southern Brazil. Pelotas has an estimated population of approximately 350,000 inhabitants, being considered a reference in the Southern region of Brazil in terms of education, since it has five higher education institutions and four large technical schools (IBGE, 2010). The UFPel is an important federal institution on the national scenario in terms of teaching, community service and research. In 2020, UFPel was ranked 40^th^ among universities in Latin America in the WHO Latin America Ranking, and among the 800-1000 best universities in the world. It has more than 160 undergraduate and graduate courses distributed in 22 academic units, and around 50 education centers of online learning courses are offered by the institution (Presidência do Brasil, 2011).

### Study design

This was a cross-sectional web-based survey conducted between July-August 2020 through a self-administered online questionnaire about the impact of the pandemic on the mental health of the academic community of UFPel. All students and staff of the University were eligible to take part in this study (n=25,220), comprising 18,814 undergraduate students, 3,781 graduate students, 1,369 academic staff and 1,256 administrative staff.

### Sample size calculation

The sample sizes for the prevalence of depression and anxiety were calculated considering a confidence interval of 95%. The sample size required given a prevalence of depression of 32% in the undergraduate population (Flesch et al., 2020), and an error margin of 5 percentage points was 618 students. For the study of anxiety, considering a prevalence of 28.4%, the largest sample size needed was 660 individuals.

### Outcome variables

Major depressive episode was evaluated using the ‘Patient Health Questionnaire-9 (PHQ-9) which assesses nine depressive symptoms according with the Diagnostic and Statistical Manual of Mental Disorders, Fourth Edition (DSM-IV): depressed mood; anhedonia, sleep disturbances; fatigue or lethargy; changes in appetite or body weight; feelings of guilt or worthlessness; difficulty in concentration; feelings of being slow or restless; and suicidal thoughts. Total score ranges from 0 to 27 points. Each question has four answer categories: 0 (not once), 1 (several days), 2 (more than half of the days), 3 (almost every day). For analysis purposes, an algorithm was calculated. The algorithm defines depression as present when the participant reports five or more symptoms, among which at least one is depressed mood or anhedonia, and that each symptom corresponds to answers 2 or 3 (‘more than half the days’ and ‘almost every day’, respectively), except for symptom 9 (suicidal thoughts), for which any value from 1 to 3 (‘less than a week’, ‘a week or more’ and ‘almost every day’, respectively) is considered as a depression symptom (Santos et al., 2013).

Anxiety disorders were assessed using the ‘Generalized Anxiety Disorder-7’ (GAD-7). This scale assesses the occurrence of seven symptoms of Generalized Anxiety Disorder in the two weeks prior to the interview. In summary, symptoms of GAD relate to feeling nervous/anxious or on edge, not being able to stop/control worrying, worrying too much, trouble relaxing, easily annoyed/irritable and feeling afraid as if something awful might happen. Total score ranges from 0 to 21 points. Each question has four answer categories: 0 (not once), 1 (several days), 2 (more than half of the days), 3 (almost every day). For analysis purposes, the cut-off 9/10 (No/Yes) was adopted (Moreno et al., 2016; Spitzer et al., 2006).

### Covariates

Gender, age group (number of years, stratified as 18-21, 22-24, 25-30, 31-41; and ≥42), skin color according to the Brazilian Census (White, Black, Mixed, East Asian, Indigenous), and family income (categorized into quintiles) were the socioeconomic and demographic variables collected.

Participants were questioned about social distancing in the period of pandemic. This information was assessed through four questions related to compliance of the authority’s guidelines for social distancing, routine of activities during the period of social distancing and perception about the importance of social distancing (Barros et al., 2020). Regarding the compliance with social distancing measures, the participant was asked ‘To what extent are you managing to follow the social distancing guidance from the health authorities, i.e., staying at home and avoiding contact with others?’. The answer was collected on a five-point scale, later combined in very little/little, some, and quite/isolated from everyone. Participant’s routine was assessed by the question ‘What have your routine activities been?’, which had as potential answers the following alternatives ‘staying home all the time’, ‘only leaving home only for essentials, such as groceries’, ‘leaving home from time to time to run errands and stretch legs’, ‘going out every day for regular activities’, and ‘out of the house all day, every day, either for work or for other regular activities’. The perception of importance of social distancing was assessed with a five-point scale related to the degree of importance attributed to social distancing by the participant. Answers were later categorized as ‘little/very little’, ‘some’, and ‘quite/extremely’. Finally, the degree of social distancing was also measured with a five-point scale and categorized into three groups: ‘not isolated/very little’, ‘some’, and ‘quite/isolated’.

Questions also evaluated the history of mental health, as following: a) regular visit to the psychiatrist/psychologist; b) time of last psychiatric and/or psychological assistance, categorized as ‘never’, ‘less than a year ago’, and ‘a year or more’; c) previous medical diagnosis of depression; d) previous medical diagnosis of anxiety.

### Data collection

The questionnaire had 65 mandatory close-ended items and was hosted online (RedCap Corporation). All eligible participants received an email through the University system with information about the survey and the questionnaire link to take part on it. The questionnaire link was also made available on the Survey official social media page on Instagram® and on Facebook®. The first page of the questionnaire contained an informed consent form. To access the questionnaire, participants had to click ‘Yes’ after the question that asked whether they agreed to participate.

Prior to data collection, a pre-test of the questionnaire was carried out assessing the understanding of the instruments used and time to complete the questionnaire. For this, four researchers were consulted. This cross-sectional study was approved by the Human Research Ethics Committee of the Federal University of Pelotas under Protocol number 4.103.085. All subjects were invited to participate and those who accepted, signed an informed consent form electronically. This study was reported according to the Strengthening the Reporting of Observational Studies in Epidemiology (STROBE) Statement and the SURGE reporting guideline (Grimshaw, 2014).

For individuals who were identified as at risk of symptoms of depression and anxiety according to the criteria previously described, the online software presented a message indicating places to seek for remote and face-to-face assistance within the University and the municipality social services networks.

### Statistical analysis

Statistical analyses were performed using Stata 16.0 (Stata Corporation, College Station, TX, USA). A descriptive analysis was performed. Distribution of variables were presented into following categories: academic or administrative staff, undergraduate and graduate students. Chi-square test for categorical variables was used.

## Results

A total of 2,822 individuals participated in this study, of which 1,637 were undergraduate students, 517 were graduate students, 229 were administrative staff and 439 were academic staff. Table 1 shows the sample representativeness in relation to the UFPel community for the variables gender, skin color and age. Women and white skin color were the majority among all subgroups (undergraduate and graduate students, academic and administrative staff) who participated in the survey. The most prevalent age category among undergraduate students was between 18 to 21 years (40.9%), and among graduate students it was between 25 to 30 years (44.9%). In both academic and administrative staff, most participants were 42 years old or more. In relation to family income, a crescent gradient was observed. Among undergraduate students, the 2^nd^ quintile of family income was the most prevalent (36.8%), among graduate students it was the 3^rd^ quintile (33.1%), 4^th^ quintile of family income among administrative staff (40.1%), and the 5^th^ quintile of family income among academic staff (72.2%) (Table 2).

**Table 1.** Sample representativeness.

|  | Undergraduate students |  |  |  | Graduate students |  |  |  | Administrative staff |  |  |  | Academic staff |  |  |  |
| --- | --- | --- | --- | --- | --- | --- | --- | --- | --- | --- | --- | --- | --- | --- | --- | --- |
|  | UFPel |  | Study sample |  | UFPel |  | Study sample |  | UFPel |  | Study sample |  | UFPel |  | Study sample |  |
|  | N | % | N | % | N | % | N | % | N | % | N | % | N | % | N | % |
| **Gender** | p<0.001 |  |  |  | p<0.001 |  |  |  | p=0.245 |  |  |  | p<0.001 |  |  |  |
| Male | 8,509 | 45.2 | 501 | 30.6 | 1,635 | 43.2 | 124 | 24.0 | 540 | 43.0 | 89 | 38.9 | 681 | 49.7 | 174 | 39.6 |
| Female | 10,305 | 54.8 | 1,136 | 69.4 | 2,146 | 56.8 | 393 | 76.0 | 716 | 57.0 | 140 | 61.1 | 688 | 50.3 | 265 | 60.4 |
| **Total** | **18,814** | **100.0** | **1,637** | **100.0** | **3,781** | **100.0** | **517** | **100.0** | **1,256** | **100.0** | **229** | **100.0** | **1,369** | **100.0** | **439** | **100.0** |
| **Skin color** | p=0.411 |  |  |  | p=0.031 |  |  |  | p= 0.691 |  |  |  | p=0.653 |  |  |  |
| White | 12,764 | 75.9 | 1,235 | 75.7 | 1,502 | 87.1 | 434 | 83.8 | 1,096 | 87.9 | 206 | 90.8 | 1,253 | 94.1 | 411 | 94.1 |
| Black | 1,824 | 10.8 | 169 | 10.4 | 109 | 6.3 | 28 | 5.4 | 74 | 5.9 | 11 | 4.9 | 18 | 1.4 | 3 | 0.7 |
| Brown | 2,092 | 12.4 | 210 | 12.9 | 99 | 5.7 | 50 | 9.7 | 71 | 5.7 | 10 | 4.4 | 56 | 4.2 | 21 | 4.8 |
| East Asian | 96 | 0.6 | 15 | 0.9 | 8 | 0.5 | 3 | 0.6 | 5 | 0.4 | 0 | 0 | 1 | 0.1 | 0 | - |
| Indigenous | 45 | 0.3 | 3 | 0.2 | 7 | 0.4 | 3 | 0.6 | 1 | 0.1 | 0 | 0 | 3 | 0.2 | 2 | 0.5 |
| **Total** | **16,821** | **100.0** | **1,632** | **100.0** | **1,725** | **100.0** | **518** | **100.0** | **1,247** | **100.0** | **227** | **100.0** | **1,331** | **100.0** | **437** | **100.0** |
| **Age** | p<0.001 |  |  |  | p<0.001 |  |  |  | p<0.001 |  |  |  | p<0.001 |  |  |  |
| 18-21 | 5,063 | 27.0 | 667 | 40.9 | 6 | 0.2 | 2 | 0.4 | 0 | - | 2 | 0.9 | 0 | - | 2 | 0.5 |
| 22-24 | 5,206 | 27.7 | 427 | 26.2 | 396 | 10.5 | 75 | 14.6 | 3 | 0.2 | 0 | - | 0 | - | 0 | - |
| 25-30 | 4,418 | 23.5 | 272 | 16.7 | 1,519 | 40.5 | 231 | 44.8 | 68 | 5.4 | 21 | 9.3 | 13 | 1.0 | 10 | 2.3 |
| 31-41 | 2,498 | 13.3 | 131 | 8.0 | 1,321 | 35.2 | 155 | 30.1 | 433 | 34.5 | 97 | 42.7 | 447 | 32.6 | 176 | 40.3 |
| ≥42 | 1,603 | 5.8 | 133 | 8.2 | 512 | 13.6 | 52 | 10.1 | 752 | 59.9 | 107 | 47.1 | 909 | 66.4 | 249 | 57.0 |
| **Total** | **18,788** | **100.0** | **1,630** | **100.0** | **3,754** | **100.0** | **515** | **100.0** | **1,256** | **100.0** | **227** | **100.0** | **1,369** | **100.0** | **437** | **100.0** |

**Table 2.** Sample characteristics.

|  | **Undergraduate**<br>**students**<br>**N (%)** | **Graduate**<br>**students**<br>**N (%)** | **Administrative**<br>**staff**<br>**N (%)** | **Academic**<br>**staff**<br>**N (%)** |
| --- | --- | --- | --- | --- |
| **Gender** |  |  |  |  |
| Male | 501 (30.6) | 124 (24.0) | 89 (38.9) | 174 (39.6) |
| Female | 1,136 (69.4) | 393 (76.0) | 140 (61.1) | 265 (60.4) |
| **Age** |  |  |  |  |
| 18-21 | 667 (40.9) | 2 (0.4) | 2 (0.9) | 2 (0.5) |
| 22-24 | 427 (26.2) | 75 (14.6) | 0 (-) | 0 (-) |
| 25-30 | 272 (16.7) | 231 (44.9) | 21 (9.3) | 10 (2.3) |
| 31-41 | 131 (8.0) | 155 (30.1) | 97 (42.7) | 176 (40.3) |
| ≥42 | 133 (8.2) | 52 (10.1) | 107 (47.1) | 249 (57.0) |
| **Skin color** |  |  |  |  |
| White | 1,235 (75.7) | 434 (83.8) | 206 (90.8) | 411 (94.1) |
| Black | 169 (10.4) | 28 (5.4) | 11 (4.9) | 3 (0.7) |
| Brown | 210 (12.9) | 50 (9.7) | 11 (4.4) | 21 (4.8) |
| East Asian | 15 (0.9) | 3 (0.6) | 0 (-) | 0 (-) |
| Indigenous | 3 (0.2) | 3 (0.6) | 0 (-) | 2 (0.5) |
| **Family income** |  |  |  |  |
| 1<sup>st</sup> quintile | 235 (16.7) | 19 (3.9) | 2 (0.9) | 1 (0.2) |
| 2<sup>nd</sup> quintile | 517 (36.8) | 151 (31.2) | 4 (1.8) | 1 (0.2) |
| 3<sup>rd</sup> quintile | 378 (26.9) | 160 (33.1) | 74 (34.1) | 7 (1.7) |
| 4<sup>th</sup> quintile | 172 (12.2) | 105 (21.7) | 87 (40.1) | 108 (25.7) |
| 5<sup>th</sup> quintile | 103 (7.3) | 49 (10.1) | 50 (23.0) | 304 (72.2) |

Prevalence of depression and anxiety were 39.2% (95% Confidence Interval (CI) 37.3-41.1) and 52.5% (95% CI 50.6-54.4), respectively. When stratified by staff and students, prevalence of depression was 49.1% (95% CI 46.5-51.6), 38.7% (95% CI 34.4-43.1), 15.5% (95% CI 11.1-21.2), and 14.7% (95% CI 11.6-18.5) among undergraduate and graduate students, administrative and academic staff, respectively. Prevalence of anxiety was 60.5% (95% CI 58.0-63.0), 53.7% (95% CI 49.3-58.1), 32.4% (95% CI 26.3-39.0), and 31.1% (95% CI 26.8-35.7) among undergraduate and graduate students, administrative and academic staffs, respectively (Figure 1).

**Figure 1.**
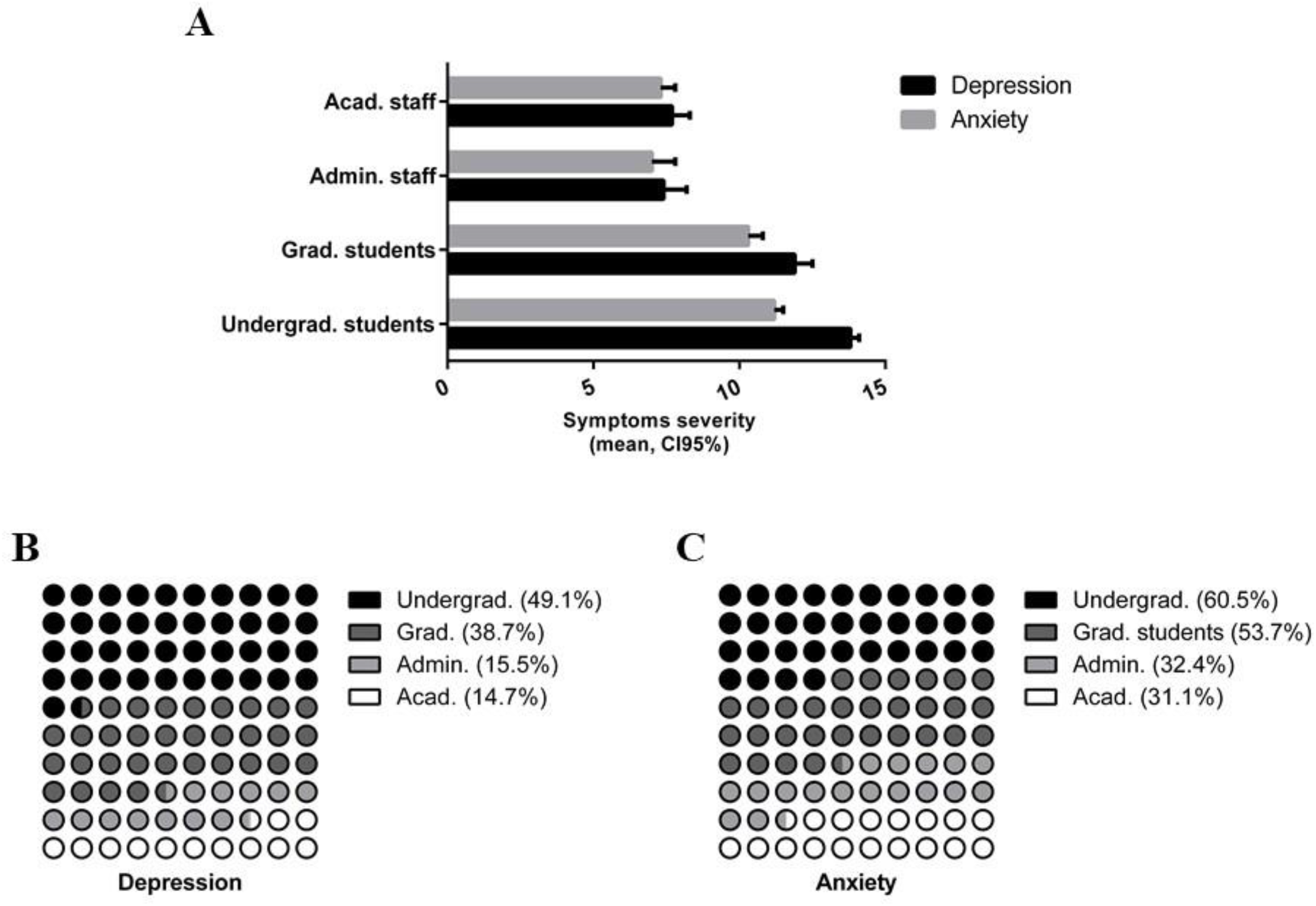
Prevalence of depression and anxiety among undergraduate and graduate students, administrative and academic staff, respectively. A) Mean of severity of symptoms; B) Percentage of depression; C) Percentage of anxiety.

Table 3 presents the association between covariates and depression. For all groups, it was observed that those who did not regularly visit the psychiatrist/psychologist, those whose time of last psychiatric and/or psychological assistance was less than a year ago and those with previous medical diagnosis of depression and/or anxiety presented higher prevalence of signals and symptoms of depression. In relation to the official recommendations about social distancing, academic staff that followed the recommendations for social distancing presented higher prevalence of depression. Considering the routine activities during the pandemic, undergraduate and graduate students who did not leave their home or left their home only for essential activities presented higher prevalence of depression.

**Table 3.**
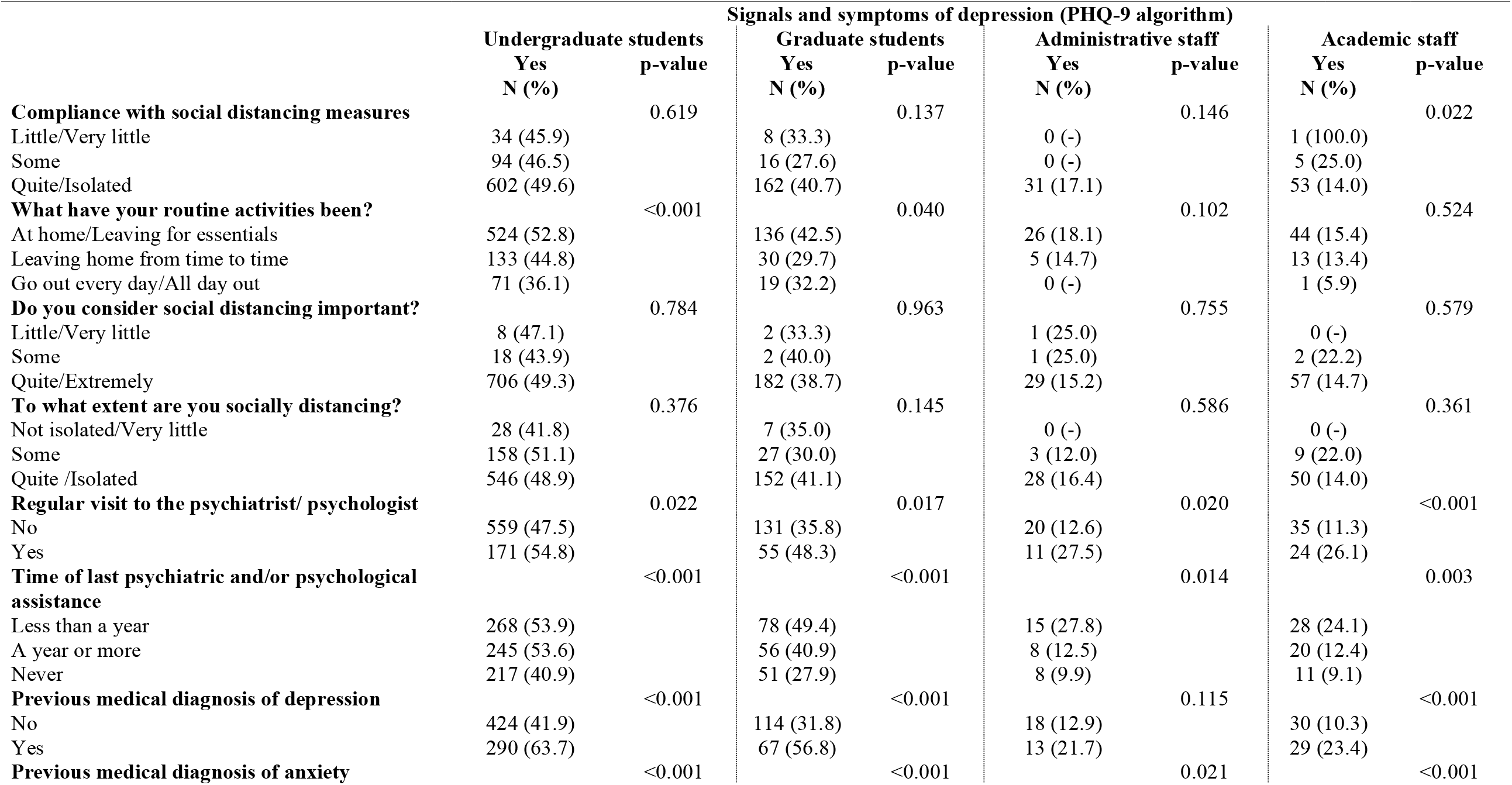
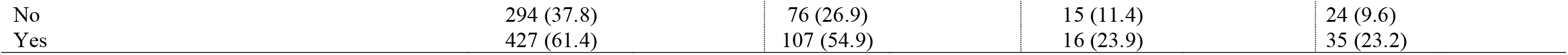
Association between sociodemographic characteristics and social distancing and depression among students and staff.

|  | Signals and symptoms of depression (PHQ-9 algorithm) |  |  |  |  |  |  |  |
| --- | --- | --- | --- | --- | --- | --- | --- | --- |
|  | Undergraduate students |  | Graduate students |  | Administrative staff |  | Academic staff |  |
|  | Yes<br>N (%) | p-value | Yes<br>N (%) | p-value | Yes<br>N (%) | p-value | Yes<br>N (%) | p-value |
| **Compliance with social distancing measures** |  | 0.619 |  | 0.137 |  | 0.146 |  | 0.022 |
| Little/Very little | 34 (45.9) |  | 8 (33.3) |  | 0 (-) |  | 1 (100.0) |  |
| Some | 94 (46.5) |  | 16 (27.6) |  | 0 (-) |  | 5 (25.0) |  |
| Quite/Isolated | 602 (49.6) |  | 162 (40.7) |  | 31 (17.1) |  | 53 (14.0) |  |
| **What have your routine activities been?** |  | <0.001 |  | 0.040 |  | 0.102 |  | 0.524 |
| At home/Leaving for essentials | 524 (52.8) |  | 136 (42.5) |  | 26 (18.1) |  | 44 (15.4) |  |
| Leaving home from time to time | 133 (44.8) |  | 30 (29.7) |  | 5 (14.7) |  | 13 (13.4) |  |
| Go out every day/All day out | 71 (36.1) |  | 19 (32.2) |  | 0 (-) |  | 1 (5.9) |  |
| **Do you consider social distancing important?** |  | 0.784 |  | 0.963 |  | 0.755 |  | 0.579 |
| Little/Very little | 8 (47.1) |  | 2 (33.3) |  | 1 (25.0) |  | 0 (-) |  |
| Some | 18 (43.9) |  | 2 (40.0) |  | 1 (25.0) |  | 2 (22.2) |  |
| Quite/Extremely | 706 (49.3) |  | 182 (38.7) |  | 29 (15.2) |  | 57 (14.7) |  |
| **To what extent are you socially distancing?** |  | 0.376 |  | 0.145 |  | 0.586 |  | 0.361 |
| Not isolated/Very little | 28 (41.8) |  | 7 (35.0) |  | 0 (-) |  | 0 (-) |  |
| Some | 158 (51.1) |  | 27 (30.0) |  | 3 (12.0) |  | 9 (22.0) |  |
| Quite /Isolated | 546 (48.9) |  | 152 (41.1) |  | 28 (16.4) |  | 50 (14.0) |  |
| **Regular visit to the psychiatrist/ psychologist** |  | 0.022 |  | 0.017 |  | 0.020 |  | <0.001 |
| No | 559 (47.5) |  | 131 (35.8) |  | 20 (12.6) |  | 35 (11.3) |  |
| Yes | 171 (54.8) |  | 55 (48.3) |  | 11 (27.5) |  | 24 (26.1) |  |
| **Time of last psychiatric and/or psychological assistance** |  | <0.001 |  | <0.001 |  | 0.014 |  | 0.003 |
| Less than a year | 268 (53.9) |  | 78 (49.4) |  | 15 (27.8) |  | 28 (24.1) |  |
| A year or more | 245 (53.6) |  | 56 (40.9) |  | 8 (12.5) |  | 20 (12.4) |  |
| Never | 217 (40.9) |  | 51 (27.9) |  | 8 (9.9) |  | 11 (9.1) |  |
| **Previous medical diagnosis of depression** |  | <0.001 |  | <0.001 |  | 0.115 |  | <0.001 |
| No | 424 (41.9) |  | 114 (31.8) |  | 18 (12.9) |  | 30 (10.3) |  |
| Yes | 290 (63.7) |  | 67 (56.8) |  | 13 (21.7) |  | 29 (23.4) |  |
| **Previous medical diagnosis of anxiety** |  | <0.001 |  | <0.001 |  | 0.021 |  | <0.001 |

|  |  |  |  |  |
| --- | --- | --- | --- | --- |
| No | 294 (37.8) | 76 (26.9) | 15 (11.4) | 24 (9.6) |
| Yes | 427 (61.4) | 107 (54.9) | 16 (23.9) | 35 (23.2) |

Table 4 shows the association between covariates and signals and symptoms of anxiety. Our findings identified that undergraduate students who never or almost never leave their home present higher prevalence of anxiety. Anxiety symptoms were also associated with non-regular visits to the psychiatrist/psychologist, to time of last psychiatric and/or psychological assistance lower than a year, and to previous medical diagnosis of depression and/or anxiety for both staff and students.

**Table 4.**
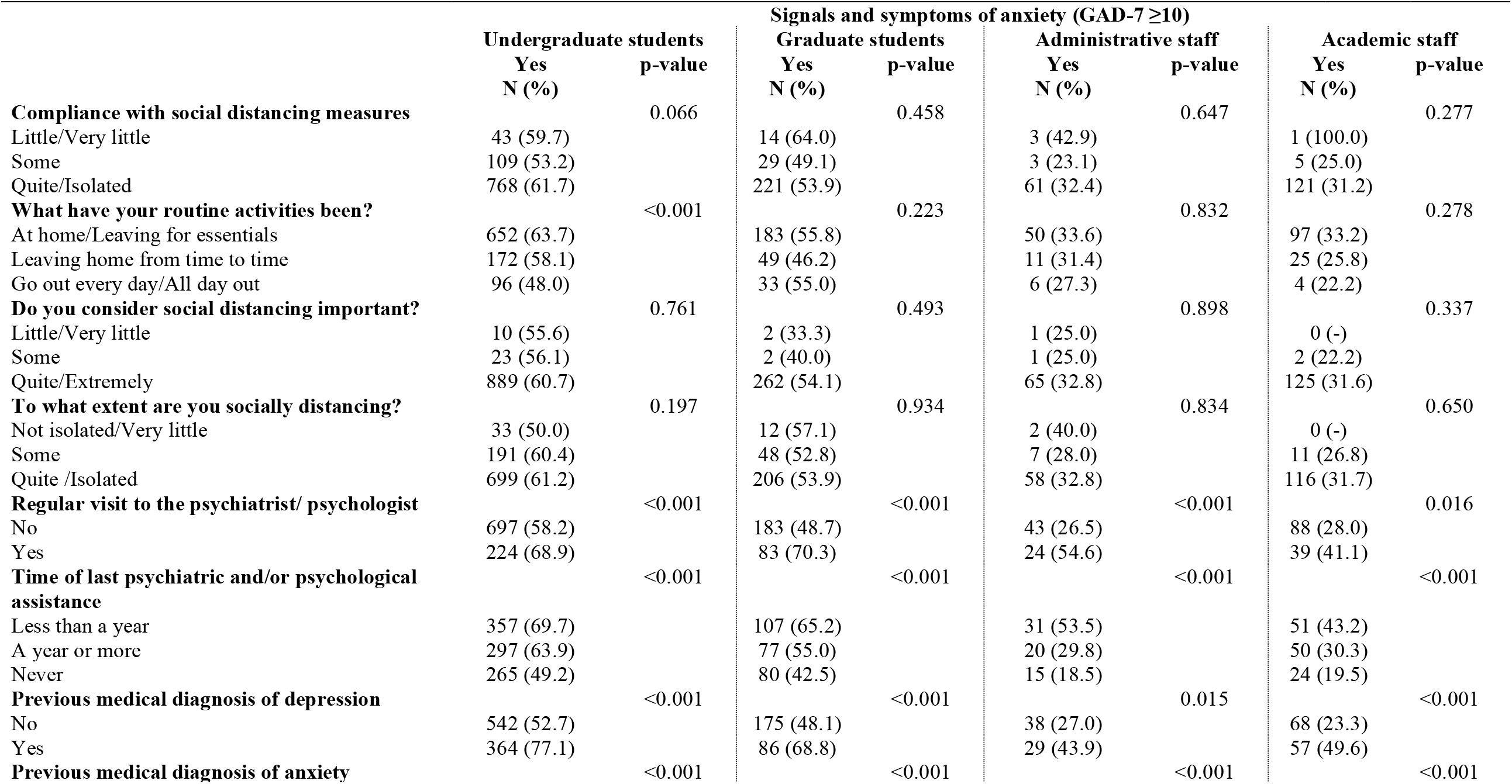
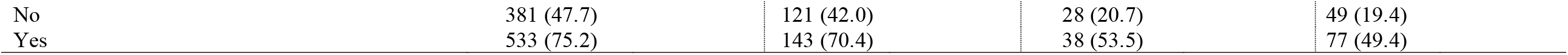
Association between sociodemographic characteristics and social distancing and anxiety among students and staff.

|  | **Signals and symptoms of anxiety (GAD-7 $\geq 10$)** |  |  |  |  |  |  |  |
| --- | --- | --- | --- | --- | --- | --- | --- | --- |
|  | **Undergraduate students** |  | **Graduate students** |  | **Administrative staff** |  | **Academic staff** |  |
|  | **Yes**<br>**N (%)** | **p-value** | **Yes**<br>**N (%)** | **p-value** | **Yes**<br>**N (%)** | **p-value** | **Yes**<br>**N (%)** | **p-value** |
| **Compliance with social distancing measures** |  | 0.066 |  | 0.458 |  | 0.647 |  | 0.277 |
| Little/Very little | 43 (59.7) |  | 14 (64.0) |  | 3 (42.9) |  | 1 (100.0) |  |
| Some | 109 (53.2) |  | 29 (49.1) |  | 3 (23.1) |  | 5 (25.0) |  |
| Quite/Isolated | 768 (61.7) |  | 221 (53.9) |  | 61 (32.4) |  | 121 (31.2) |  |
| **What have your routine activities been?** |  | <0.001 |  | 0.223 |  | 0.832 |  | 0.278 |
| At home/Leaving for essentials | 652 (63.7) |  | 183 (55.8) |  | 50 (33.6) |  | 97 (33.2) |  |
| Leaving home from time to time | 172 (58.1) |  | 49 (46.2) |  | 11 (31.4) |  | 25 (25.8) |  |
| Go out every day/All day out | 96 (48.0) |  | 33 (55.0) |  | 6 (27.3) |  | 4 (22.2) |  |
| **Do you consider social distancing important?** |  | 0.761 |  | 0.493 |  | 0.898 |  | 0.337 |
| Little/Very little | 10 (55.6) |  | 2 (33.3) |  | 1 (25.0) |  | 0 (-) |  |
| Some | 23 (56.1) |  | 2 (40.0) |  | 1 (25.0) |  | 2 (22.2) |  |
| Quite/Extremely | 889 (60.7) |  | 262 (54.1) |  | 65 (32.8) |  | 125 (31.6) |  |
| **To what extent are you socially distancing?** |  | 0.197 |  | 0.934 |  | 0.834 |  | 0.650 |
| Not isolated/Very little | 33 (50.0) |  | 12 (57.1) |  | 2 (40.0) |  | 0 (-) |  |
| Some | 191 (60.4) |  | 48 (52.8) |  | 7 (28.0) |  | 11 (26.8) |  |
| Quite /Isolated | 699 (61.2) |  | 206 (53.9) |  | 58 (32.8) |  | 116 (31.7) |  |
| **Regular visit to the psychiatrist/ psychologist** |  | <0.001 |  | <0.001 |  | <0.001 |  | 0.016 |
| No | 697 (58.2) |  | 183 (48.7) |  | 43 (26.5) |  | 88 (28.0) |  |
| Yes | 224 (68.9) |  | 83 (70.3) |  | 24 (54.6) |  | 39 (41.1) |  |
| **Time of last psychiatric and/or psychological assistance** |  | <0.001 |  | <0.001 |  | <0.001 |  | <0.001 |
| Less than a year | 357 (69.7) |  | 107 (65.2) |  | 31 (53.5) |  | 51 (43.2) |  |
| A year or more | 297 (63.9) |  | 77 (55.0) |  | 20 (29.8) |  | 50 (30.3) |  |
| Never | 265 (49.2) |  | 80 (42.5) |  | 15 (18.5) |  | 24 (19.5) |  |
| **Previous medical diagnosis of depression** |  | <0.001 |  | <0.001 |  | 0.015 |  | <0.001 |
| No | 542 (52.7) |  | 175 (48.1) |  | 38 (27.0) |  | 68 (23.3) |  |
| Yes | 364 (77.1) |  | 86 (68.8) |  | 29 (43.9) |  | 57 (49.6) |  |
| **Previous medical diagnosis of anxiety** |  | <0.001 |  | <0.001 |  | <0.001 |  | <0.001 |

|  |  |  |  |  |
| --- | --- | --- | --- | --- |
| No | 381 (47.7) | 121 (42.0) | 28 (20.7) | 49 (19.4) |
| Yes | 533 (75.2) | 143 (70.4) | 38 (53.5) | 77 (49.4) |

## Discussion

The prevalence of depression and anxiety among undergraduate students was 49.1% and 60.5%, respectively. Rates of both psychopathologies were higher among those students who had ever been diagnosed with depression/anxiety and who reported visiting healthcare providers in the previous year. In relation to social distancing, among some but not all subgroups higher prevalence of anxiety and depression mental illness was associated to strictly following the authority’s guidelines for social distancing and to not leaving the house routinely. These associations were not pronounced in all groups, but it may indicate the lack of face-to-face social interaction during pandemic as a cause of depression and anxiety.

Large-scale, disruptive crises such as a pandemic have profound short and long-term impacts on population mental health, including depression, anxiety, post-traumatic stress disorder, psychological distress, and stress (Taquet et al., 2021; Xiong et al., 2020). Alarming prevalence of depression and anxiety were observed for all individuals in this study, with figures considerably higher in comparison to a systematic review and meta-analysis of mental health consequences of COVID-19, which identified overall prevalence of depression and anxiety of 31.4% and 31.9%, respectively (Wu et al., 2021). Not surprisingly, an association between mental health issues and previous medical diagnosis of depression and/or anxiety was identified. The scientific literature suggests the COVID-19 pandemic may impact on the mental health of the general population and worse the psychiatric symptoms of those with pre-existing psychiatric disorders (Shigemura et al., 2020; Vindegaard and Benros, 2020), and this finding was corroborated by the present study.

Our findings identified that COVID-19 is taking a larger toll on the mental health of university students in comparison to staff: almost half of students were identified to have depression, and 60.0% of them presented anxiety symptoms. Four main reasons can be hypothesized to this higher mental health burden among university students. Firstly, staff at Federal Universities in Brazil have permanent positions and standard wages. While the economic impacts of the pandemic are substantial and there are well documented consequences of income instability to mental health (Allen et al., 2014), it is expected that this problem will be less concerning for those who have stable employment and salary, which were not affected by the economic crisis triggered by the Pandemic. Secondly, students are doing a course and have uncertainties about their future, differently than staff in permanent positions. The pandemic affected their way of learning, moving from face-to-face lectures to e-learning, and significantly impacted their expected graduation dates, which may lead to mental health issues. Additionally, it is a common practice in Brazil for students to move intercity or interstate to attend Federal Universities, and the lack of social support in a new city may make these individuals more vulnerable to the mental health effects of a stressful period such as the COVID-19 pandemic. Finally, students are usually younger than staff. A study evaluating the epidemic of a highly infectious equine influenza in Australia identified age as a factor associated with the level of psychological distress, with those individuals aged 16 to 24 showing the highest levels of mental health impact. A systematic review on the impact of COVID-19 on mental health also showed student status and younger age group to be risk factors associated with mental distress during the pandemic (Xiong et al., 2020). While evidence shows that older people are more susceptible to the physical effects of Covid-19, it seems the long-term mental health burden may be more dangerous to the younger groups.

Social distancing, quarantine and isolation are recommended by public health authorities for the prevention of the transmission of infectious diseases, such as COVID-19 (Brooks et al., 2020). However, social isolation is an established risk factor for mental health, and social support and connections are of critical importance during major health events, including quarantine and isolation (Hossain et al., 2020). Social distancing is the reduction of social contacts, while quarantine is the separation of people potentially exposed to a contagious disease (Brooks et al., 2020). Although with different levels of severity, both are used to minimize the spread of an infectious disease. In this study, students and academic staff who reported following the recommendations for social distancing were identified to present more symptoms of depression and anxiety. Before the introduction of vaccines, social distancing, mask use and hand sanitation were the only effective measures to prevent the uncontrolled spreading of the virus, preventing the health systems of collapsing and saving lives (Bedford et al., 2020). Despite these well-known benefits of social distancing and quarantine as a public health measure, some conditions should be observed to minimize its mental health impact. Conditions identified as stressors during quarantine include its duration, the fear of becoming infected or transmitting the virus to others, feelings of frustration and boredom, inadequate information, and inadequate supplies, both general and medical. While social distant, the fear of the infection may predispose individuals to be hypervigilant for symptoms, which may increase their levels of fear and anxiety. Also, the sense of isolation can be distressing and may have psychological impacts. Quarantine may have a considerable, long-term psychological effects for those affected, and measures should be taken to reduce its impact (Brooks et al., 2020).

The full impact of the COVID-19 pandemic on mental health is still unknown, but the evidence that a psychological effect of quarantine may still be detected months or years later is worrisome, and suggests the need to ensure, even during the period, effective governmental and individual efforts to reduce mental health effects. Inefficiently receiving information from public health authorities can contribute as a stressor to the population, and one of the actions that can be taken is to promote clear, accurate, and up-to-date communication aimed at promoting a good understanding of the disease in question, reducing insecurities, and increasing awareness. In addition, those with a mental illness history may be more likely to experience psychological distress after experiencing any disaster-related trauma, so they need to be under close watchful eye for any additional support during this stressful pandemic period. The pandemic and related quarantine also appear to have a larger impact on health care workers than non-health care workers, and the society and governments should be responsive to the mental health needs of health workers (Brooks et al., 2020). Additionally, the basic needs, including food, water, and basic medical supplies of those quarantined should be met by the government, to reduce the mental burden of an already stressful experience. Finally, the health system needs to be prepared to deal with the long-term mental health effects of a societal traumatic event such as a pandemic.

The strengths of this study include the instruments used to assess mental health outcomes and the University’s official support to the survey. The PHQ-9 and the GAD-7, used to assess depression and anxiety, respectively, are widely used, validated instruments, therefore granting reliability to our estimates. Additionally, the survey was officially supported by the University, by inviting the academic community to take part on the study via e-mail and by advertising it on the University website and official social media accounts. This is also one of the first studies to be developed in relation to mental health in the University during the Pandemic period and the results shed some light in relation to the effects of pandemic in the academic community and the need to tackle the problem.

The main concern is with the representativeness of the sample, especially in relation to gender and age. The fact that the study sample was younger than the University’s community may reflect the data collection process, carried out completely online. In addition, the response rate was relatively low, which can also reflect the limitations of online self-administered questionnaires. Also, the rates of full completion of online questionnaires are lower for web-based interviews that the figures for face-to-face data collections (Heiervang and Goodman, 2011). Another point worth mention is that individuals with more severe mental health issues may be less likely to engage in web-based surveys, leading to underestimated prevalence of these problems. Finally, the cross-sectional nature of our data limits the evaluation of temporality or the mental health monitoring over different moments of the pandemic. Also, it is worth mentioning that the questionnaire was applied when the online activities had just started. Academic activities were suspended on March 16^th^, and students and academic staff had more than 3 months without in person or online teaching process., while the University was moving to the remote teaching. This scenario could increase the risk for depression and anxiety symptoms or loneliness among University students (Sundarasen et al., 2020).

In conclusion, COVID-19 is a global pandemic that may shape the mental health of a whole generation. The overall levels of anxiety and depression during the COVID-19 pandemic are alarming, and the levels among University students are specially concerning. Individuals with previous medical diagnosis of mental illness and those practicing social isolation appear to have higher prevalence of depression and anxiety symptoms.

## Data Availability

The database for this study is not available on an external database. However, they can be made available if requested.

